# Blood-based immunophenotyping of T cell profiles in patients with neurodegenerative disorders

**DOI:** 10.1101/2025.09.19.25336156

**Authors:** Frederika Malichova, Peter Swann, Stacey L Kigar, Natalia Savinykh Yarkoni, Julia Goddard, Leonidas Chouliaras, Ajenthan Surendranathan, Lorinda Turner, George Savulich, Richard Bevan-Jones, Nicholas J Ashton, Kaj Blennow, Henrik Zetterberg, Edward Needham, Joanne Jones, William A McEwan, James B Rowe, John T O’Brien, Maura Malpetti

**Affiliations:** University of Cambridge Department of Clinical Neurosciences and Cambridge University Hospitals NHS Trust, Cambridge, United Kingdom; Department of Psychiatry, University of Cambridge, Cambridge, United Kingdom; Department of Medicine, University Cambridge, Cambridge, United Kingdom; UK Dementia Research Institute at University of Cambridge, Cambridge CB2 0XY, UK; Specialty Dementia and Frailty Service, Essex Partnership University NHS Foundation Trust, Epping, UK; University College London Hospitals NHS Foundation Trust; Department of Psychiatry and Neurochemistry, University of Gothenburg, Gothenburg, Sweden; Banner Alzheimer’s Institute and University of Arizona, Phoenix, AZ, USA; Banner Sun Health Research Institute, Sun City, AZ 85351, USA; Clinical Neurochemistry Laboratory, Sahlgrenska University Hospital, Mölndal, S-431 80, Sweden; Clinical Neurochemistry Laboratory, Sahlgrenska University Hospital, Mölndal, Sweden; Department of Neurodegenerative Disease, UCL Institute of Neurology, Queen Square, London, UK; UK Dementia Research Institute at UCL, London, UK; Hong Kong Center for Neurodegenerative Diseases, InnoHK, Hong Kong, China; Wisconsin Alzheimer’s Disease Research Center, University of Wisconsin School of Medicine and Public Health, University of Wisconsin-Madison, Madison, WI, USA; Medical Research Council Cognition and Brain Sciences Unit, Cambridge, United Kingdom

## Abstract

**Background:** There is increasing evidence for the role of central and peripheral inflammation across neurodegenerative disorders, with animal models and post-mortem studies identifying T-cell infiltration in the brain associated with pathology and neurodegeneration. Peripheral T-cell changes have been measured in Alzheimer’s disease (AD) with conflicting results and limited characterization. This study examines blood-based T-cell profiles across a range of neurodegenerative dementias including AD, dementia with Lewy bodies (DLB), frontotemporal dementia (FTD), corticobasal syndrome (CBS), progressive supranuclear palsy (PSP), and aged-matched healthy controls, testing for associations with dementia-relevant plasma biomarkers and clinical outcomes.

**Methods:** Freshly prepared peripheral blood mononuclear cells (PBMCs) from 174 participants (AD=20, DLB=24, FTD=19, CBS=18, PSP=58, controls=35) were studied using a flow-cytometry panel designed to analyse major T-cell subpopulations, including memory and T-helper subtypes. Neurodegeneration-relevant biomarkers (p-tau217, p-tau231, GFAP, NFL, and A-beta42/40) were measured in plasma samples. T-cell populations were compared between groups and in association with biomarkers, and principal components analysis (PCA) was used to identify T-cell profiles and their association with dementia-relevant biomarkers in diagnostic classification and survival prediction.

**Results:** There was a significant reduction in the proportion of CD3+ cells in patients with DLB compared to other diagnostic groups, and an increase in relative Th1/17 cell levels in patients with AD and FTD compared to controls. This increase in Th1/17 cells correlated with NfL and GFAP plasma levels in FTD. PCA identified four components primarily representing CD4+ memory cell population subsets. There was an increase in Th1/17 and Th17 effector memory profiles in AD and FTD. These cellular profiles were limited in diagnostic classification compared to p-tau217 or NfL, but the profile of increased naïve CD4+ cells with decreased Th1 effector memory cells was associated with mortality across all diseases.

**Conclusions:** This study provides evidence for T-cell dysregulation and diagnosis-specific profiles in neurodegenerative diseases, further establishing adaptive immunity as a key contributor to disease heterogeneity. Although plasma biomarkers such as NfL and p-tau217 exhibit superior diagnostic accuracy for clinical classification, peripheral T-cell signatures were associated with survival outcomes across diagnostic groups, highlighting their promise for prognostic applications and disease monitoring. The characterisation of T-cell populations across neurodegenerative conditions may inform target development and patient stratification for new interventional trials.

## BACKGROUND

The primary roles of the immune system are healing and defence against pathogens and harmful agents including bacteria, viruses, toxins, or damaged cells (1). Broadly, it can be classified into two branches: the innate and the adaptive immune system. The innate immune system is the first line of defence against pathogens and consists of cells including monocytes and neutrophils. The adaptive immune system is antigen-specific and has long lived memory cells, allowing rapid recognition of previously encountered antigens. The adaptive immune system consists of two major cell populations, the B and T-cells (2).

Neuroinflammation is established as a hallmark across neurodegenerative diseases(3). Microglial activation, as measured by positron emission tomography (PET) of the translocator protein (TSPO), is associated with disease progression and clinical severity in patients with neurodegenerative dementias (4–7). Increasingly, studies are exploring the role of peripheral immune system (8–11). Systemic inflammation has been shown to exacerbate neuropathology in animal models of neurodegeneration and is associated with disease progression in clinical cohorts (12). This may be mediated by T-cell migration across the blood-brain/blood-cerebrospinal fluid barrier (13–15). T-cell infiltration promoting astrocytosis, microgliosis and neurodegeneration has been reported in mouse models of tauopathy(16,17) and the presence of T-cells in human post-mortem brain tissue from patients with Alzheimer’s disease (AD), frontotemporal lobar degeneration (FTLD), and dementia with Lewy bodies (DLB) (18–22). The role of T-cells in neurodegeneration is not clear, with studies finding associations with neuronal loss (23), whilst others highlight a potential protective role of specific T-cell subsets (24).

Although the peripheral adaptive immune system, and specifically T-cell populations, plays a role in neurodegeneration, further studies are needed particularly comparing AD and non-AD dementias to identify whether T-cell changes are disease-specific or transdiagnostic across neurodegeneration. Characterisation of these cells could lead to clinically relevant blood-based biomarkers to support target identification and patient stratification for immune-targeting therapies. Conflicting evidence on the proportion of CD4+ and CD8+ T-cell proportions has been reported in AD and related animal models, however very few looked at this across multiple neurodegenerative diagnoses (25–29). There are studies of peripheral T-cell immunophenotyping, predominantly in AD (30), only few studies have compared this across a range of neurodegenerative diseases, in association with plasma neurodegenerative biomarkers, or explored associations with survival.

This study therefore aims to characterise T-cell profiles in blood from people with neurodegenerative diseases causing dementia, including AD, DLB, frontotemporal dementia (FTD), corticobasal syndrome (CBS) and progressive supranuclear palsy (PSP). Using flow cytometry, we immunophenotyped freshly prepared peripheral blood mononuclear cells (PBMCs) to characterise and quantify T-cell sub-populations. We tested three hypotheses: firstly, that there are differences in the proportion of major T-cell populations (CD3+, CD4+, CD8+, memory subsets and T-helper lineages) across neurodegenerative disorders that correlate with other neurodegenerative biomarkers, secondly that T-cell profiles provide useful information to classify neurodegenerative diagnosis, and finally that T-cell profiles are associated with survival.

## METHODS

### Participants

Patients were recruited from clinics for cognitive and movement disorders at the Cambridge University Hospitals NHS Trust as well as collaborating local psychiatry and neurology services. We included participants with AD (comprising people who met criteria for Mild Cognitive Impairment (31) with evidence of AD pathology or standardised criteria for AD (32) (n=20), CBS (n=18) (33), DLB (34), FTD (n=19; 13 behavioural variant FTD (bvFTD) and 6 primary progressive aphasia (PPA)) (35,36), and probable or possible PSP (n=58, predominantly Richardson’s syndrome) (37). Age-matched healthy volunteers (n=35) were included as the control group, with MMSE > 26/30, absence of memory symptoms, no signs of dementia, or other significant medical illnesses (38). Exclusion criteria for both patient and healthy control cohorts included major concurrent psychiatric illness, other severe physical illness, or a history of other significant neurological illness. For participants with mental capacity, all gave written informed consent according to the Declaration of Helsinki. For participants who did not have the mental capacity to consent, the consultee process was followed. The study protocol was approved by the NIHR National Research Ethic Service Committee and East of England (Cambridge Central; REC Ref: 13/EE/0104; 07/Q0102/3).

Participants underwent a standardized clinical assessment with the Addenbrooke’s Cognitive Examination (39) (ACE-R), which represents a global measure of cognition scored out of 100. The ACE-R can be subdivided into five domains: Attention and Orientation (18/100), Memory (26/100), Fluency (14/100), Language (26/100), and Visuospatial abilities (16/100). Survival data were collected for all participants up to and including the 27^th^ November 2024 (the census date).

### Blood collection and flow cytometry

At baseline, 18 ml of blood was drawn into EDTA blood tubes and analyzed at the NIHR Cambridge BRC Cell Phenotyping Hub. Wherever possible, samples were processed within 2h (over 98% of samples). Blood was layered onto sterile Ficoll (Cytiva, Cat#: 17144003) for peripheral blood mononuclear cell (PBMC) isolation by a technician blind to group status. Two aliquots containing ∼1 x10^6 PBMCs were labelled using live-dead stains and antibodies for CD3, CD4, CD8, CXCR3, CD45RA, CCR7, and CCR6. At the end of staining, cells were washed, and the data were acquired on live (i.e., non-fixed) cells with a BD LSR Fortessa instrument. Cell classifications were determined via manual gating in FlowJo (BD) by individuals blind to the participant group according to standards recommended for the Human Immunology Project (40) using an agreed gating strategy (**Supplementary Figure 1**). For clarity of communication, we defined CD3+ cells as T-cells, CCR7+CD4+5RA+ cells as naïve, CCR7+CD4+5RA-cells as central memory (CM), CCR7-CD4+5RA-cells as effector memory (EM), and CCR7-CD4+5RA+ cells as terminally differentiated effector memory cells re-expressing CD4+5RA (EMRA/TEMRA). We also defined CXCR3+CCR6-CD4+ T cells as Th1 cells, CXCR3-CCR6-CD4+ T cells as Th2 cells, CXCR3-CCR6+ CD4+ T cells as Th17 cells and CXCR3+CCR6+CD4+ T cells as Th1/17 cells, including only memory populations in these subsets (by removing naïve cells defined as CCR7+ CD45+) and recognising that these are enriched rather than pure populations of Th subtypes. The median number of live events recorded for each participant was 387,208, and the median for the CD4+ memory total was 65628. All subpopulations had a median number of cells greater than 100. Whilst the fewest cells were in CD4+ TEMRA subpopulation, all but three of the participants (98%) had greater than 100 CD4+ TEMRA cells measured. The median cell numbers in each gate are shown in **Supplementary Table 1**.

### Plasma biomarkers relevant to neurodegenerative diseases

In a sub-cohort of 94 participants (AD=18, CBS=12, DLB=20, FTD=13, PSP=31), plasma samples stored at -70°C were used for further analyses at the Clinical Neurochemistry Laboratory in Mölndal (Sweden). Plasma samples were thawed on wet ice, centrifuged at 500×[g for 5 min at 4°C. Calibrators (neat) and samples (plasma: 1:4 dilution) were measured in duplicates. The plasma assays performed were the Quanterix Simoa Human Neurology 4-Plex E assay (measuring Aβ40, Aβ42, GFAP and NfL, Quanterix, Billerica, MA), the p-tau217 ALZpath assay measuring p-tau217 of the human tau protein associated with AD, and p-tau231 with an in-house Simoa assay as previously described (41,42). Samples were analysed at the same time using the same batch of reagents. A four-parameter logistic curve fit data reduction method was used to generate a calibration curve. Two control samples of known concentration of the protein of interest (high-control and low-control) were included as quality control. Intra-assay coefficients of variation were below 10%. The mean plasma biomarker concentrations for each group are shown in **Supplementary Table 2**.

### Statistical analysis

Statistical analyses were carried out in R for frequentist tests and JASP for Bayesian tests. Differences between sex were compared with the Chi-squared test. Group differences on continuous variables were compared Kruskal-Wallis Chi-squared test, followed by Dunn’s post-hoc test with Benjamin-Hochberg corrections for multiple comparisons. Where there were differences in demographic covariates, linear models were used with the relevant covariates with the T-cell population proportion as the outcome and group as a predictor. Bayesian ANOVA was used in JASP using ranks, and Bayes Factor compared to the null model reported. A Bayes Factor (BF10) between 1 and 3 is regarded as ‘mild’, between 3 and 10 as ‘moderate’, between 10 and 30 as ‘strong’ and above 30 as ‘very strong’ evidence for H1 over H0.

Principal components analysis (PCA) was performed using the *prcomp* R function, selecting components with an adjusted eigenvalue >1 from Horn’s parallel analysis, and applying a varimax rotation to the retained components. Individual component scores were extracted and used as variables in multinomial and then logistic regression. Correlations were performed with Spearman’s rank correlation coefficient. Cox proportional hazard models were performed in R with the time between blood draw and death (or census date if the patient was still alive) as the outcome, and T-cell component scores as predictors.

### Demographics and baseline cognition

One hundred and thirty-nine participants with neurodegenerative dementias were recruited and thirty-five cognitive unimpaired controls. Overall, there was no significant difference in age, but there were differences in sex (DLB group proportionally more male than CBS group) As expected, all dementia groups had lower cognitive scores, as measured by the ACE-R, compared to controls p<0.001, and AD and FTD lower than PSP, t=-3.46, p=0.009, t=-2.94, p=0.043 respectively, post-hoc Dunn tests).

**Table 1:** Demographics and baseline cognition: 1: Mean (SD); n (%), 2: Wilcoxon rank sum test; Pearson’s Chi-squared test, 3: Kruskal-Wallis rank sum test; Pearson’s Chi-squared test. M=male, F=female.

|  | Control<br>N = 35<sup>1</sup> | Dementia<br>(all)<br>N = 139 | AD<br>N = 20<sup>1</sup> | CBS<br>N = 18<sup>1</sup> | FTD<br>N = 19<sup>1</sup> | PSP<br>N = 58<sup>1</sup> | DLB<br>N = 24<sup>1</sup> | All dementia<br>vs control<sup>2</sup> | Individual group<br>differences<sup>3</sup> |
| --- | --- | --- | --- | --- | --- | --- | --- | --- | --- |
| Age | 69.0<br>(6.4) | 71.3 (7.5) | 71.0<br>(6.0) | 69.3<br>(8.9) | 69.3<br>(7.6) | 71.7<br>(7.1) | 73.4<br>(8.0) | W=1966,<br>p=0.080 | H = 8.7, p = 0.12 |
| Sex |  |  |  |  |  |  |  | X<sup>2</sup>=0.42,<br>p=0.5 | X<sup>2</sup>=14.3, p =<br>0.014 |
| F | 13<br>(37%) | 60 (43%) | 8<br>(40%) | 12<br>(67%) | 6<br>(32%) | 30<br>(52%) | 4<br>(17%) |  |  |
| M | 22<br>(63%) | 79 (57%) | 12<br>(60%) | 6<br>(33%) | 13<br>(68%) | 28<br>(48%) | 20<br>(83%) |  |  |
| ACER | 95.9<br>(3.4) | 74.3 (17.2) | 66.2<br>(14.5) | 75.1<br>(17.7) | 67.9<br>(23.8) | 79.6<br>(14.1) | 73.3<br>(15.9) | W=4389,<br>p<0.001 | H = 75.6, p<br><0.001 |

### Group differences in major T-cell populations

There were significant group differences in CD3+ cells, as a proportion of live PBMCs (H=14.4, p=0.013, BF_10_=2.35), with participants in the DLB group having significantly lower CD3+ levels than all other disease groups (Dunn’s post-hoc test with Benjamin-Hochberg corrections, DLB vs AD: Z =-2.80, pFDR=0.029, BF_10_=11.36; DLB vs FTD: Z=-2.60, pFDR=0.035, BF_10_=6.05; DLB vs PSP: Z=-2.75, pFDR=0.029, BF_10_=5.02; DLB vs CBS: Z=-3.30, pFDR=0.013, BF_10_=35.47), with moderate to very strong evidence from Bayesian ANOVAs (see **Figure 1A**). However, the identified difference in CD3+ cells between patients with DLB and controls did not survive correction for multiple comparisons (Z=-2.00, uncorrected p=0.046, corrected p=0.14) with the Bayes-Factor (BF_10_=1.66) suggesting mild evidence in favour of a difference. These results remained the same when controlling for age, and sex, in a linear model with CD3+ cell proportion as the main outcome and diagnostic group as the main predictor (DLB vs AD: p=0.01; DLB vs FTD: p=0.050, DLB vs PSP: p=0.037; DLB vs CBS: p=0.017). This could represent either a decrease in CD3+ cell levels, or an increase in other cell types leading to the proportional decrease.

**Figure 1:**
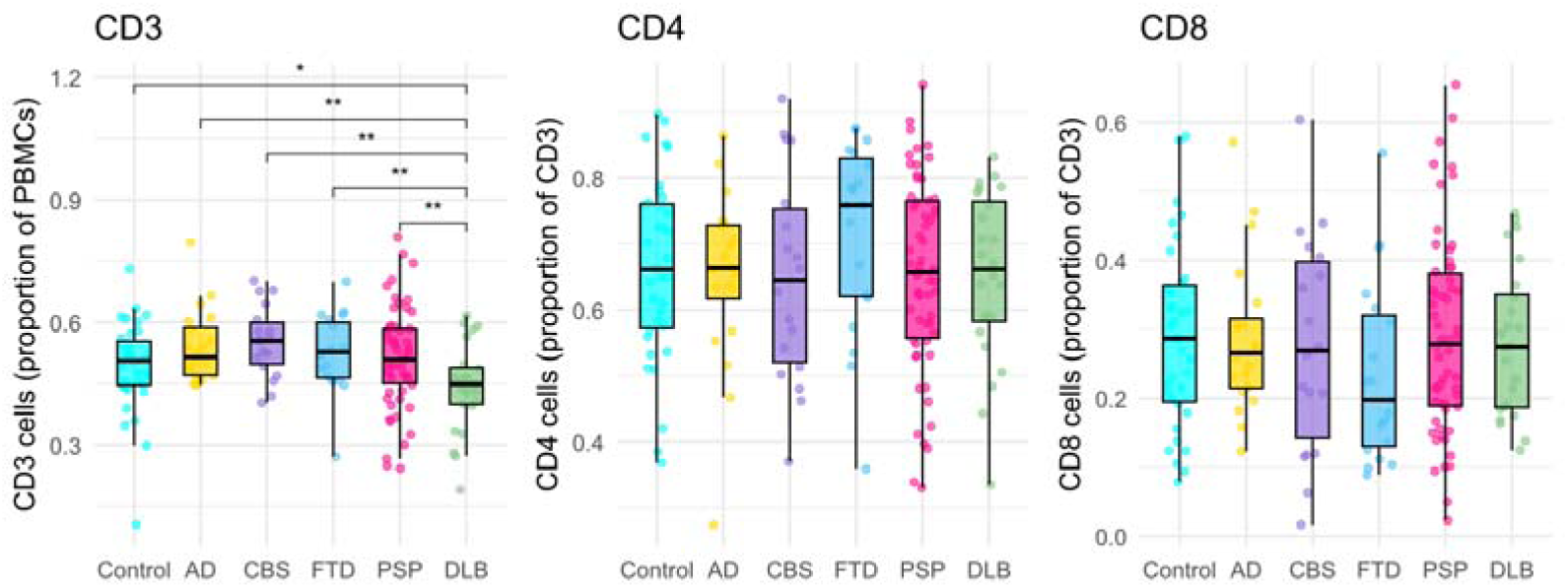
Group differences in CD3+, CD4+ and CD8+ T-cell populations. Box and dot plots display CD3+ (left), CD4+ (middle), and CD8+ (right) cell proportions between groups. Each bar represents a different group (AD-Alzheimer’s disease, CBS-corticobasal syndrome, FTD-frontotemporal dementia, PSP-progressive supranuclear palsy, DLB-dementia with Lewy bodies), with dots representing individuals within that group. Double asterisks indicate significance with FDR correction and single asterisk indicates uncorrected (p<0.05) significance from Dunn’s post-hoc test with Benjamin-Hochberg correction following a significant Kruskal-Wallis test.

There were no significant group differences in CD4+ cells (H=3.9, p=0.56, BF_10_ = 0.043) or CD8+ cells (H=3.36, p=0.64, BF_10_=0.035) as a proportion of CD3+ cells (Kruskal-Wallis test, Bayesian ANOVA) (**Figure 1**). Likewise, there were no significant group differences in the CD4+/CD8+ ratio (H=3.45, p=0.63, BF_10_ =0.085).

#### Group differences in T-cell subpopulations

Between groups, as a proportion of total memory CD4+ T-cells, there was no significant difference in Th1 (H=5.18, p=0.39, BF_10_ = 0.104), Th2 (H=2.77, p=0.73, BF_10_ =0.037), or Th17 (H=4.31, p=51, BF_10_ =0.050) cells but a significant difference in Th1/17 cells was seen (H=11.38, p=0.044, BF_10_ =1.125, Kruskal-Wallis, Bayesian ANOVA) with increases in AD vs controls (p=0.008, pFDR=0.04), FTD vs controls (p=0.01, pFDR=0.042), and AD and FTD compared to PSP (AD: p=0.008, pFDR= 0.042 and FTD: p=0.01, pFDR=0.042), as indicated by the Dunn’s post-hoc test. The linear model with Th1/17 cells as outcome variable, groups as main predictor, and age and sex as covariates, identified significant differences in patients with AD (t=2.75, p=0.007) and FTD (t=2.20, p=0.029) as compared to controls (**Figure 2**). To ensure this was not driven by individuals with low CD4+ memory pools, we ran a sensitivity analysis excluding those with less than 1000 cells (one participant from the PSP group) and results remained significant (Kruskal-Wallis; p=0.038, AD vs control; pFDR=0.039, FTD vs control; pFDR=0.039, AD vs PSP; pFDR=0.039, FTD vs PSP; pFDR=0.039).

**Figure 2:**
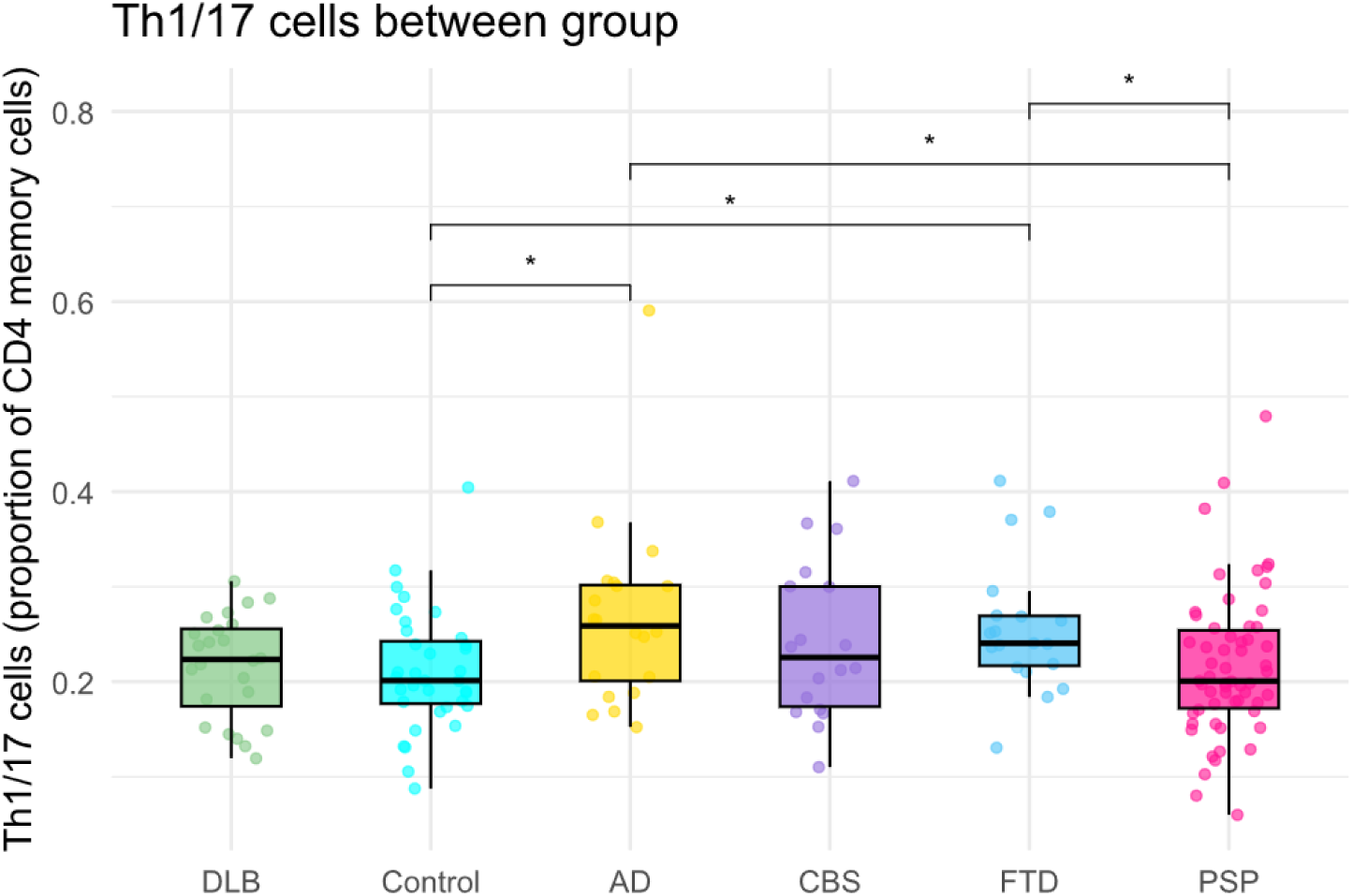
Group differences in Th1/17 cells. Box and dot plots displaying Th1/17 cells as a proportion of total memory cells. Each bar represents a different group, with dots representing individuals within that group (AD-Alzheimer’s disease, CBS-corticobasal syndrome, FTD-frontotemporal dementia, PSP-progressive supranuclear palsy, DLB-dementia with Lewy bodies). * indicates (p<0.05) significance from Dunn’s post-hoc test with Benjamin-Hochberg correction following a significant Kruskal-Wallis test.

There were no group differences in naïve (H=4.5 p=0.47, BF_10_ = 0.072), CM (H=6.1 p=0.29, BF_10_ = 0.493), EM (H=5.0 p=0.41, BF_10_ = 0.074), or TEMRA (H=8.9 p=0.11, BF_10_ = 0.146) cells as a proportion of overall CD4+ cells (Kruskal-Wallis test, Bayesian ANOVA). When including age and sex in linear models, we identified a decrease in naïve CD4+ cells in PSP (t=-2.12, p=0.036). Regarding CD8+ cells, there was similarly no overall group differences in naïve (H=7.6 p=0.18, BF_10_ = 0.187), CM (X^2^=7.5 p=0.18, BF_10_ = 0.308), EM (X^2^=8.0 p=0.16, BF_10_ = 0.169), or TEMRA (X^2^=8.5 p=0.13, BF_10_ = 0.146) cells as a proportion of overall CD8+ cells (Kruskal-Wallis test, Bayesian ANOVA). However, the linear models including age and sex as covariates identified a reduction in naïve CD8+ cells in PSP (t=-1.98, p=0.049), and in CD8+ CM cells in both PSP (t=-2.25, p=0.026) and DLB (t=-2.57, p=0.011) compared to controls.

### Association of T-cell populations with plasma dementia-relevant biomarkers

Next, we explored the relationship between T-cell populations and plasma biomarkers with linear models and partial correlations, correcting for age and sex. We focused on the cell populations identified as showing different proportions between disease groups and controls (CD3+ cells in DLB; Th1/17 cells in AD and FTD).

The proportion of CD3+ cells in patients with DLB was not a significant predictor of p-tau217 (t=-0.13, p=0.90), p-tau231 (-0.35, p=0.71), GFAP (t=0.71, p=0.48), or NfL (t=-1.29, p=0.20) in linear models controlling for age and sex.

In patients with AD, Th1/17 cell proportion was not a significant predictor of p-tau217 (t=1.66, p=0.11), p-tau231(t=0.76, p=0.46), GFAP (t=1.40, p=0.17), or NfL (t=0.52, p=0.61), in linear models controlling for age and sex.

Similarly, in patients with FTD Th1/17 cell proportions were not a significant predictor of p-tau217 (t=1.46, p=0.14) or p-tau231 (t=1.25, p=0.22), however they were significant predictors of NfL (t=4.38, p=0.0001) and GFAP (t=2.90, p=0.0064). Age (t=3.22, p=0.0028) and sex (t=2.53, p=0.016) were also significant predictors of NfL, and age was a significant predictor of GFAP (t=2.79, p=0.0086). There was a moderate positive partial correlation between Th1/17 cells and NfL in FTD, controlling for age and sex (Spearman’s rho=0.57, p<0.001), and between Th1/17 cells and GFAP, controlling for age and sex (Spearman’s rho=0.45, p=0.0055) (**Figure 3**).

**Figure 3:**
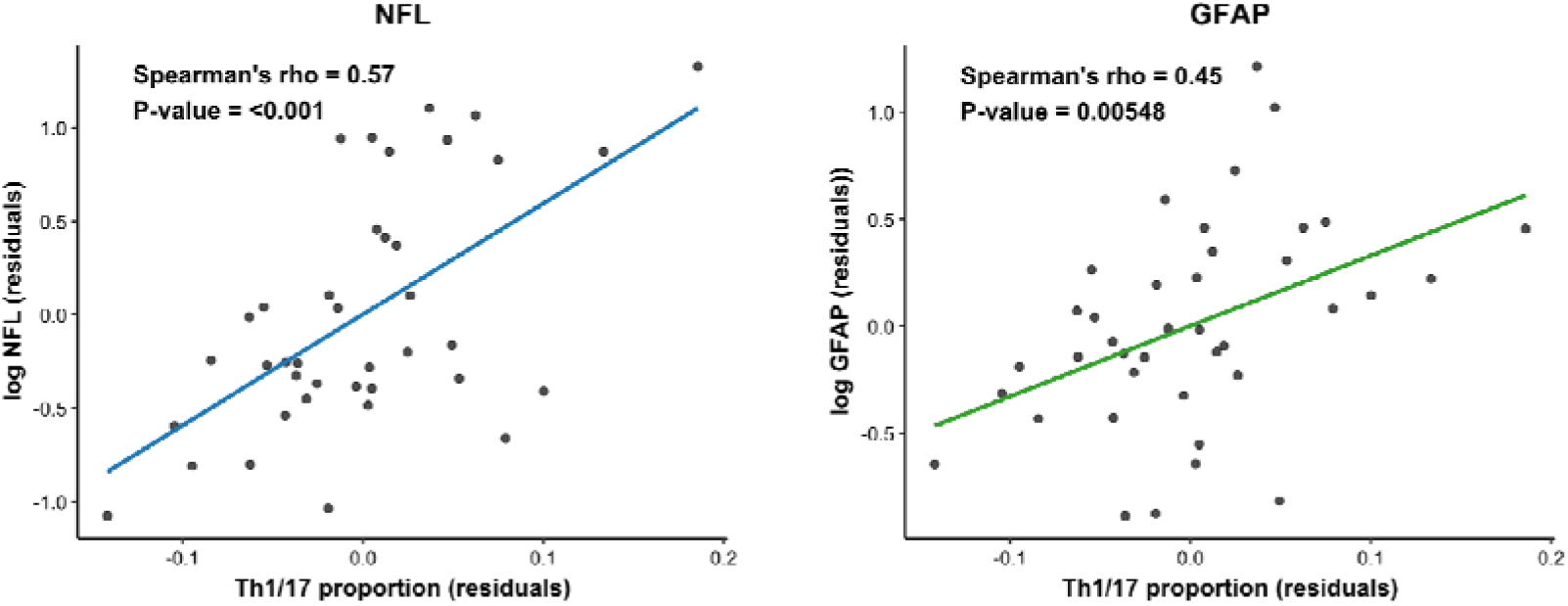
Associations of Th1/17 memory cell proportions as a percent of total memory CD4+ cells with NfL (left) and GFAP (right). Plots show the residuals of log(NfL) and Th1/17 cell proportion from a linear model including age and sex (as significantly associated covariates), and the residuals of log(GFAP) correcting for age.

### PCA identifies T-cell profiles across neurodegenerative diagnosis

Finally, we included 17 subpopulations, both CD4+ and CD8+, as a proportion of total live CD3+ cells in a PCA (**Supplementary Table 3**). The overall MSA was 0.54 and Bartlett’s Test of Sphericity significant, suggesting the data were adequate for PCA. Results of Horn’s parallel analysis suggested including four components, which in total explained 60% of the data. The variable loadings of component 2 and component 3 were predominantly negative so both variable loadings for these components were multiplied by -1 for interpretability. The component loadings reflected the hierarchical gating with each component broadly representing a CD4+ memory cell phenotype (component 1 = CM, component 2 = TEMRA, component 3 = EM, and component 4 = naïve cells), with component 3 also predominantly weighted on the CCR6+ populations, i.e., Th1/17 and Th17 cells (Figure 4A). Next, we used multinomial regression to see if scaled individual scores for these components were significant predictors of diagnostic groups, with Wald Chi squared test to assess significance of individual predictors. Using group as the outcome, with age and sex as covariates, we identified component 3 (representing CCR6+ EM CD4+ populations) was a significant predictor of FTD (odds ratio = 2.06, CI=1.11-3.83, p=0.02) and AD (odds ratio = 1.96, CI=1.06-3.63, p=0.032) (Figure 4B). As the CCR6+ EM populations in the PCA clustered together with increases in FTD and AD rather than following our nominal separation into Th1/17 and Th17 cells, we tested to see if CCR6+ (Th1/17 + Th17) memory cell proportions were increased in these groups as a proportion of the total memory pool. Total CCR6+ CD4+ memory cells were proportionally increased in FTD (W=203, p=0.018) but not significantly in AD (W=239, p=0.053). Conversely, CCR6+ CD4+ EM cells were proportionally increased in AD (W=195, p=0.0061), but not FTD (W=240, p=0.095). Component 2, representing CD4+ TEMRA populations, was increased in PSP compared to controls (W=1298, p=0.025) but did not reach statistical significance in the multinomial model as a predictor of diagnostic group (odds ratio = 1.97, CI= 0.97-4.01, p=0.062).

**Figure 4:**
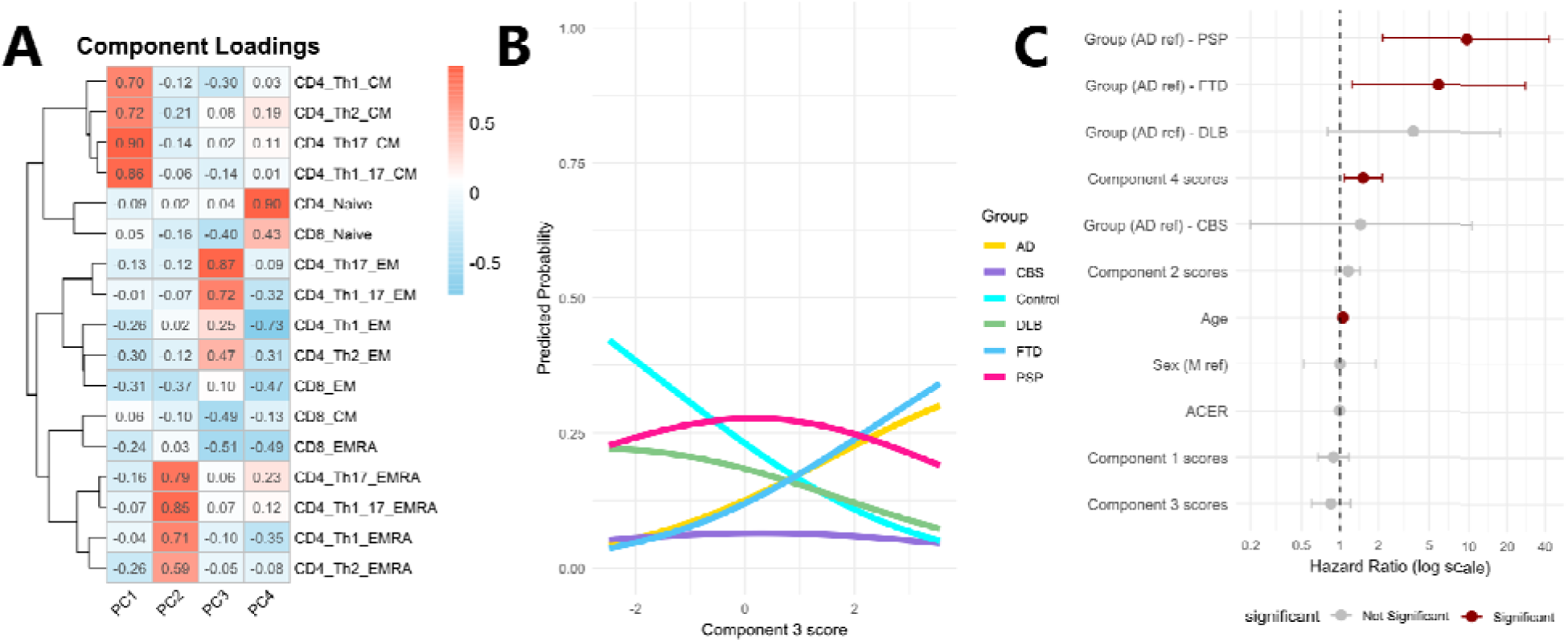
T-cell profiles across diagnosis are predictive of survival. A: Heatmap showing the loadings of four principal components identified by PCA. Each row represents a T-cell subpopulation and each column a component, with the colour showing the loading of each T-cell population in that component. B: Marginal effects plot showing the predicted probability of each diagnostic group controlling for age and sex. This shows that increased component 3 scores are associated with an increased predicted probability of AD and FTD compared to other diagnoses. C: Forrest plot showing the results of the Cox proportional hazards model including the principal component scores representing T-cell profiles, with diagnosis, age, sex, and ACE-R as covariates. This shows that higher component 4 scores are associated with increased mortality, along with age and a diagnosis of progressive supranuclear palsy (PSP) or frontotemporal dementia (FTD) (compared to the reference group, Alzheimer’s disease (AD)).

### T-cell profiles in diagnostic classification

To test the usefulness of these T-cell profiles as tools for differential diagnoses, we compared the component scores in logistic regression models with (i) each diagnostic group vs controls, and then (ii) each diagnostic group vs other diseases. We considered age and sex as covariates and as a basic model, compared to those including individual scores for each component as predictor. Component 3 scores improved classification of AD and FTD compared to controls but were not as accurate as plasma p-tau217 and NfL respectively. Receiver operating curves (ROC) for these models are shown in **Supplementary Figure 2**.

### T-cell profiles predict survival

Finally, we assessed the association of the four T-cell components with mortality using Cox proportional hazards models controlling for age, sex, diagnostic group, and total ACE-R score. We included 136 participants with dementia (53 deceased, 83 alive on date of census, 27^th^ November 2024) and did not include controls.

Components 1, 2 and 3 did not show significant prognostic effects. Increased component 4 scores, representing higher proportions of CD4+ naïve cells and lower proportions of Th1 EM cells, were associated with increased mortality (HR=1.52, p=0.017). Compared to the reference group of AD, PSP (HR=9.63, p=0.003) and FTD (OR=5.90, p=0.025) were associated with increased mortality, as was age (HR=1.05, p=0.025). To assess how this compared to other neurodegenerative biomarkers, we tested component 4 scores in a model with NfL and p-tau217, where it remained a significant predictor of mortality (HR=1.87, p=0.006) (Figure 4C).

## Discussion

In this study, a large cohort comprising 174 patients with neurodegenerative diseases and age- and sex-matched healthy controls was investigated to characterize T-cell immune profiles using comprehensive and data-driven methodologies on peripheral blood mononuclear cells. Leveraging advanced techniques for immune phenotyping, we identified marked differences in peripheral T-cell subpopulations between diagnostic groups, which were significantly associated with clinical diagnosis, survival outcomes, and established pathology biomarkers. These findings provide robust support for the mounting evidence that T-cell immunity is intricately involved not only in the pathogenesis of AD but also in non-AD dementias, underscoring a critical role for adaptive immune alterations across diverse neurodegenerative syndromes.

First, we identified group differences in CD3+ cell counting, but not in CD4+ and CD8+ total cell counts. Specifically, patients with DLB showed a lower proportion of CD3+ cell (relative to live PBMCs) than the rest of the diagnostic groups and controls. Notably, while some prior studies in patients with DLB have reported a reduction in blood CD4+ T-cell counts (11), the decrease in overall CD3+ T-cell proportion, which reflects reduced total T-cell levels, may indicate more global T-cell alterations in DLB that extend beyond the helper subset. Previous evidence suggests that T-cells are recruited into the brain in DLB (18), with CD4+ T-cells identified in the brain and CSF in patients with DLB (28). This migration may explain the observed peripheral depletion, as T-cells re-localise from the blood to the central nervous system in response to neuroinflammatory signals. These findings align with prior reports that patients with DLB have abnormal T-cell trafficking and increased evidence of central immune activation (19), supporting the hypothesis that peripheral immunological shifts mirror central immune processes. The reduction in circulating CD3+ T-cells in DLB adds further support to suggestions that altered immune homeostasis is a disease-relevant process in Lewy body disorders.

Second, we found that increased proportions of Th1/17 memory cells in AD and FTD, with an effector memory phenotype. This expands on previous research focused on single neurodegenerative diseases compared to controls. Increases in CCR6+ and T-cell memory populations have previously been described in AD (43–45), PBMCs stimulated with amyloid beta shift towards a Th17 phenotype in AD compared to controls (46), and Th1 and Th17 cells accelerated memory impairments and neuroinflammation in animal models of AD (47) and a meta-analysis has also shown increases in these Th1- and Th17-associated cytokines in AD, including TNF-alpha and IL-6 (48). Effector memory cells have also been identified in the CSF in patients with AD and FTD (49) and accumulating in the brain in AD (50). There have been fewer studies of the T-cell profiles in FTD (51), however increases in Th1 and Th17 cells have been associated with disease severity in clinical ALS (52,53) and animal models (54). Recent work has shown peripheral cytokines that are functionally linked to Th1 and Th17 responses (including TNF-alpha, IL-12, IL17A, and IL-6) are also associated with lower survival rates in patients with FTD and related disorders (4). Th1/17 cell counts were correlated with plasma NfL and GFAP in FTD, suggesting an association between peripheral T-cells, astrocyte activation and neurodegeneration. In contrast, there was no association between Th1/17 cells and plasma p-tau217 in AD. This could suggest that the immune response in AD is a separate pathological process rather than a direct response to amyloid or tau, and potentially an alternative or complementary treatment target to be explored. Few studies have explored the links between T-cells and dementia biomarkers, with one finding a blunted Th17 response associated with cognition, NfL, and GFAP in women with MCI (55) and another finding associations with CD8+ T-EMRA cells, NfL and GFAP in a community cohort of ageing and mild dementia (56).

Next, our data-driven approach identified two components that were related to clinical symptoms. Firstly, component 3 was mainly loaded onto CD4+ effective memory cells and was overrepresented in patients with AD and FTD compared to other diagnoses. This result is supported by recent studies highlighting the involvement of CD4+ T-cell subsets in neurodegenerative diseases. Notably, effector memory CD4+ T cells have been found in higher proportions in patients with AD than in healthy controls and linked with cognitive deficits (51,57,58), suggesting their potential role in the disease course and dementia-related neuroinflammation. The pathogenic role of EM CD4+ cells in accelerating memory impairment, amyloid deposition, microglia activation, and neuroinflammation in AD has further been demonstrated in animal models (47). Mechanistically, effector memory CD4+ cells are producers of pro-inflammatory cytokines and have the capacity to cross the blood– brain barrier during inflammatory states, interacting with microglia and amplifying neuroinflammatory cascades. This mechanism has been implicated not only in AD but this works potentially expands this into other neurodegenerative diseases such as FTD, suggesting a shared vulnerability driven by peripheral immune dysfunction.

Another important T-cell profile emerged from our data was Component 4, representing higher proportions of CD4+ naïve cells and lower proportions of Th1 EM cells. Higher scores on this component were associated with increased mortality rates across diagnoses. This study does not explore the mechanisms for this association or identify casual linkage, however there are several hypotheses that may explain this finding. Enhanced immune cell migration into tissues, including the brain, would reduce their proportion in the periphery, and T-cell infiltration into the brain has been associated with tau pathology and neurodegeneration (23). Alternatively, T-cell activation has been proposed to be beneficial in neurodegeneration. Studies have explored enhancing the peripheral immune response in AD (59), and T-cell exhaustion and association with loss of function may be a therapeutic target (60). Finally, lack of effective peripheral immune populations may also put individuals at higher risk of infection (55), a leading cause of mortality in dementia (61,62).

Despite some commonalities across studies, there are wide variations in the literature on T cells in patients with dementia (30). For example, in our cohort we did not identify a reduction in CD4+ cells, in contrast to previous reports in patients with AD cases (26), or the associated shift in the CD4+/CD8+ ratio, which has been previously described in blood of PSP patients (63). Other studies, however, have also not replicated these findings(64), identifying shifts towards more later differentiated CD4 T-cells with senescence markers (65), or increases in CD4 T-EMRA cells (26) in AD compared to controls. The inconsistent findings between studies may reflect differences in cohorts (26), the panel design (65) and antibody choice (51,65), with some studies on freshly collected PBMCs and others on those stored in liquid nitrogen (26). Studies with larger cohorts based on clinical and biomarker criteria across dementia groups will allow a greater understanding of the relationship between peripheral immune changes and disease progression, and stratification by disease stage or co-pathology (such as AD co-pathology in CBS or DLB).

This study has several limitations. Patients were recruited and included in groups defined on the basis of clinical diagnostic criteria, and amyloid biomarker positivity confirmation for patients with AD. Future studies would benefit from applying similar methods in cohorts of patients with pathology-confirmed diagnoses. This study characterised immunophenotypes at a single timepoint in individual participants. Immune processes in AD, similarly to other diseases, have been proposed to peak early in disease (9), and functional genomics have linked an initial proinflammatory response to tau pathology to later immunosuppression (66). Future studies with longitudinal biosampling are required to identify how these immune changes progress over individual disease courses. Our study focused on peripheral blood mononuclear cells and did not measure infiltration of T-cells into the brain or CSF, which could reveal complementary information and evidence for the role of T-cells in neurodegeneration. Combining peripheral measures of T-cell profiles with CSF or neuroimaging, and ultimately pathological measures of inflammation will lead to a more comprehensive understanding of the immune landscape across neurodegeneration. Importantly, our immunophenotyping panel relied on surface markers rather than intracellular staining to classify the major T-cell subpopulations. For example, in our study Th2 cells were defined by lack of CXCR3 and CCR6 without CCR4, while Th17 cells were defined as CXCR3-CCR6+ and may contain a very small population of Th22 cells which would be CCR10+. The lack of CD56 in our panel meant that natural killer T-cells (NKTs) were not separated from CD8+ T-cells. Recent advances in flow cytometry has enabled the characterisation of larger panels improving the classification of subpopulations, and with high parameter spectral flow cytometry leading to the detection of novel immune subpopulations in neurodegeneration (28). Future efforts to replicate these findings using larger cell phenotyping panels across a range of neurodegenerative dementias, integrating single cell RNA sequencing and T-cell receptor repertoire analysis (67), will be important to gain further insight in T-cell profiles in dementias. In addition, immunophenotyping was performed on fresh samples on the day of collection, which prevents cryopreservation-related cell death and changes in expression markers associated with freezing and thawing (68). However, this could introduce batch effects. Future studies could include internal controls to measure this with greater precision. Finally, we did not measure several confounders that could influence T-cell phenotypes such as APOE (69) and for viral infection, notably herpes simplex virus type I (HSV-1) (70) and cytomegalovirus (CMV) (56).

Overall, our findings highlight significant alterations in peripheral T-cell subpopulations in neurodegenerative diseases that correlate with distinct clinical diagnoses and plasma biomarkers of neurodegeneration. While plasma NfL and p-tau217 demonstrated superior diagnostic accuracy, the immune profile characterized by higher proportions of CD4+ naïve cells and lower proportions of Th1 effector memory cells was strongly associated with survival outcomes. This suggests that T-cell immunophenotyping may offer valuable prognostic information and serve as a potential biomarker to monitor treatment response in immunomodulatory trials. With the advent of novel T-cell-targeting therapies emerging for neurodegenerative diseases, an improved understanding of disease-specific T-cell profiles and the development of robust immune biomarkers will be crucial for patient stratification and optimizing clinical trial design.

## Data Availability

Anonymized processed data can be shared upon request with the corresponding author. Raw data may also be requested but are likely to be subject to a data transfer agreement with restrictions required to comply with participant consent and data protection regulations.

## Acknowledgements

This study was co-funded by the Dementias Platform UK and Medical Research Council (MC_UU_00030/14; MR/T033371/1); Race Against Dementia Alzheimer’s Research UK (ARUK-RADF2021A-010); the Wellcome trust (103838; 220258); the Cambridge University Centre for Parkinson-Plus (RG95450); the National Institute for Health Research (NIHR) Cambridge Biomedical Research Centre (BRC-1215-20014; NIHR203312: the views expressed are those of the authors and not necessarily those of the NIHR or the Department of Health and Social Care); the Progressive Supranuclear Palsy Association (PSPA2022/SMALL GRANTS002); the Addenbrookes Charitable Trust (Ref: 900380). We thank our participant volunteers for their participation in this study, thank the National Institute for Health Research (NIHR) Cambridge BioResource centre staff, and the research nurses for their contribution, the staff at the NIHR Cambridge Cell Phenotyping Hub at the university of Cambridge and the East Anglia Dementias and Neurodegenerative Diseases Research Network (DeNDRoN) for help with subject recruitment. EN is supported by the Medical Research Council and Brain Research UK.

