## Supplementary for "Blood-based immunophenotyping of T cell profiles in patients with neurodegenerative disorders"

Supplementary material:

Supplementary figure 1: Gating strategy

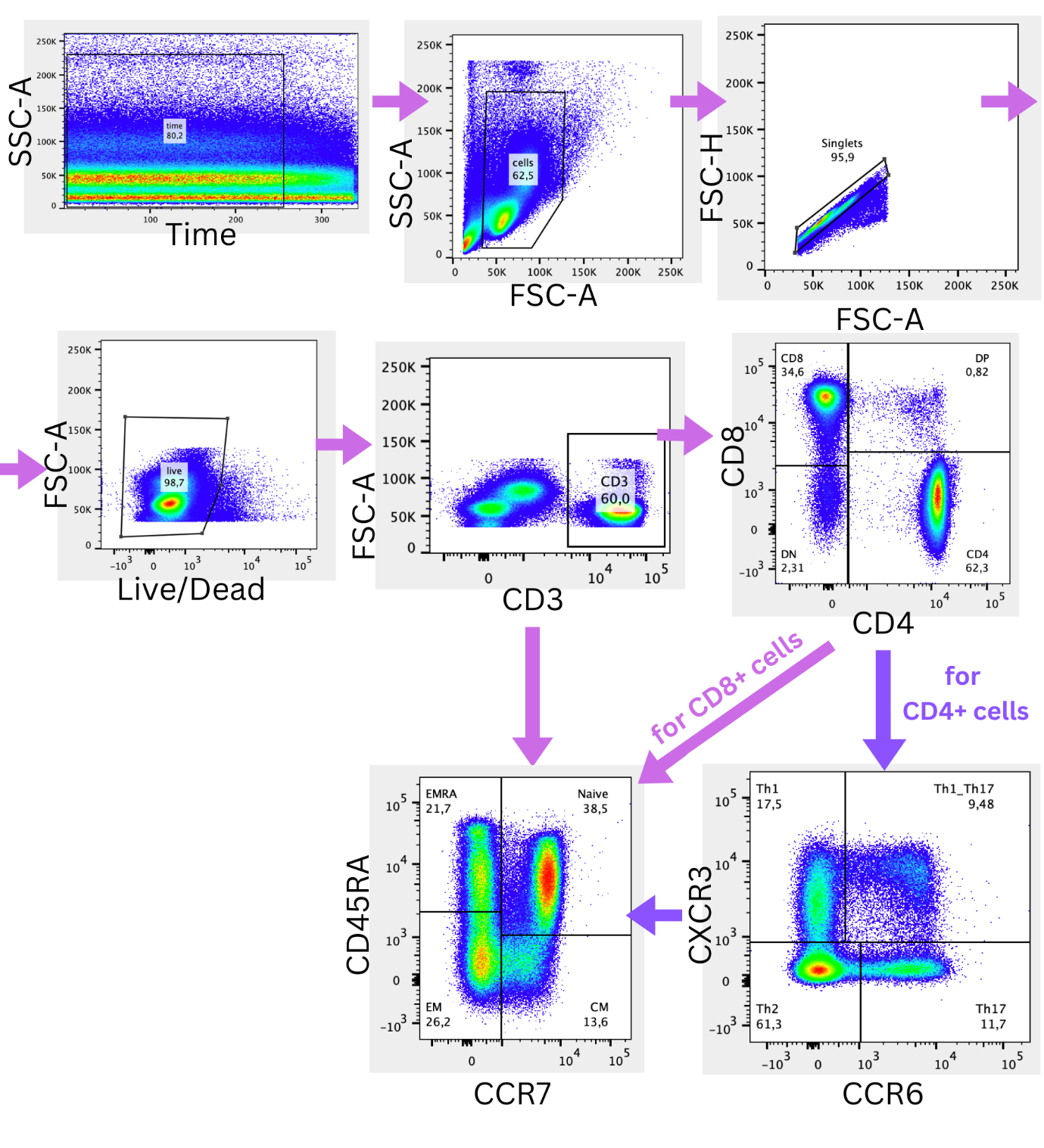

Supplementary figure 1: Gating strategy. Note that for CD4+ ‘Th’ cells, as provisionally indicated on the bivariate plots here, naïve cells were removed post-data acquisition (e.g., Th1 cells would be defined as the counts from the CD4+CXCR3+CCR6- gate minus counts from the derivative CD45RA+CCR7+ gate). In working with these data, we found this gating strategy improved interrater reliability in a subset of samples.  Thus, all data presented on Th cells in the main paper are memory cell populations, consistent with the differentiation of T helper subsets after naïve cell exposure to antigen.

Supplementary table 1: Cell numbers in each gate: median + interquartile range for each population across all patient samples

| Variable | Median | IQR (Q1-Q3) |
| --- | --- | --- |
| time_cells_singlets_live | 387,208.0 | 246404 - 532848.5 |
| time_cells_singlets_live_cd3 | 197,963.5 | 113800.25 - 257557 |
| time_cells_singlets_live_cd3_cd4 | 123,670.5 | 71899.5 - 172377.5 |
| time_cells_singlets_live_cd3_cd4_th1 | 23,532.5 | 13783 - 34399.75 |
| time_cells_singlets_live_cd3_cd4_th1_cm | 4,974.5 | 2205 - 8008.75 |
| time_cells_singlets_live_cd3_cd4_th1_em | 10,006.0 | 4406 - 17660 |
| time_cells_singlets_live_cd3_cd4_th1_emra | 1,172.0 | 581.25 - 2487.5 |
| time_cells_singlets_live_cd3_cd4_th17 | 15,803.5 | 9583.75 - 24988 |
| time_cells_singlets_live_cd3_cd4_th17_cm | 5,924.0 | 2716.5 - 10206.25 |
| time_cells_singlets_live_cd3_cd4_th17_em | 6,924.5 | 3716.75 - 12336 |
| time_cells_singlets_live_cd3_cd4_th17_emra | 173.0 | 69.25 - 404.5 |
| time_cells_singlets_live_cd3_cd4_th1_17 | 16,004.0 | 8162 - 24249.75 |
| time_cells_singlets_live_cd3_cd4_th1_17_cm | 3,503.0 | 1413.25 - 6787.5 |
| time_cells_singlets_live_cd3_cd4_th1_17_em | 10,123.0 | 4671.75 - 17390.5 |
| time_cells_singlets_live_cd3_cd4_th1_17_emra | 219.5 | 98.25 - 522.25 |
| time_cells_singlets_live_cd3_cd4_th2 | 58,210.5 | 31193.5 - 90443.5 |
| time_cells_singlets_live_cd3_cd4_th2_cm | 9,217.0 | 4402.75 - 14178.5 |
| time_cells_singlets_live_cd3_cd4_th2_em | 4,772.5 | 2349.25 - 9176.25 |
| time_cells_singlets_live_cd3_cd4_th2_emra | 762.5 | 336.25 - 1800 |
| time_cells_singlets_live_cd3_cd8 | 46,830.0 | 23940.25 - 74368.75 |
| time_cells_singlets_live_cd3_cd8_cm | 1,075.5 | 337.5 - 3261.75 |
| time_cells_singlets_live_cd3_cd8_em | 11,232.0 | 3681 - 22624.75 |
| time_cells_singlets_live_cd3_cd8_emra | 19,026.5 | 8543.25 - 47958.75 |
| time_cells_singlets_live_cd3_cd8_naive | 5,249.5 | 2620.75 - 9883.5 |

Supplementary table 2: Plasma biomarker results split by group, all measurements in pg/mL.

| Biomarker | Control | DLB | AD | CBS | FTD | PSP | Kruskal-Wallis Test (χ², p-value) |
| --- | --- | --- | --- | --- | --- | --- | --- |
| pTau217 | 0.32 ± 0.22 | 0.68 ± 0.4 | 0.91 ± 0.38 | 0.54 ± 0.47 | 0.41 ± 0.22 | 0.48 ± 0.47 | 38.05, p=< 0.001 |
| pTau231 | 6.07 ± 2.28 | 11.03 ± 7.04 | 11.37 ± 5.4 | 8.8 ± 4 | 8.06 ± 5.09 | 9.12 ± 4.51 | 23.5, p=< 0.001 |
| GFAP | 105.71 ± 55.08 | 173.26 ± 91.95 | 204.09 ± 106.3 | 206.94 ± 92.99 | 167.25 ± 101.59 | 170.22 ± 127.68 | 18.89, p=0.002 |
| NFL | 18.08 ± 6.76 | 38.08 ± 17.83 | 33.06 ± 13.87 | 61.78 ± 45.22 | 58.72 ± 25.51 | 46.22 ± 24.79 | 50.43, p=< 0.001 |

Supplementary table 3:

Table showing the cell populations included in the PCA

| Population name | Markers |
| --- | --- |
| CD4_Naive | CD3+, CD4+, CXCR3-, CCR6-, CD45RA+, CCR7+ |
| CD4 Th1 CM | CD3+, CD4+, CXCR3+, CCR6-, CD45RA-, CCR7+ |
| CD4 Th2 CM | CD3+, CD4+, CXCR3-, CCR6-, CD45RA-, CCR7+ |
| CD4 Th17 CM | CD3+, CD4+, CXCR3-, CCR6+, CD45RA-, CCR7+ |
| CD4 Th1_17 CM | CD3+, CD4+, CXCR3+, CCR6+, CD45RA-, CCR7+ |
| CD4 Th1 EM | CD3+, CD4+, CXCR3+, CCR6-, CD45RA-, CCR7- |
| CD4 Th2 EM | CD3+, CD4+, CXCR3-, CCR6-, CD45RA-, CCR7- |
| CD4 Th17 EM | CD3+, CD4+, CXCR3-, CCR6+, CD45RA-, CCR7- |
| CD4 Th1_17 EM | CD3+, CD4+, CXCR3+, CCR6+, CD45RA-, CCR7- |
| CD4 Th1 EMRA | CD3+, CD4+, CXCR3+, CCR6-, CD45RA+, CCR7- |
| CD4 Th2 EMRA | CD3+, CD4+, CXCR3-, CCR6-, CD45RA+, CCR7- |
| CD4 Th17 EMRA | CD3+, CD4+, CXCR3-, CCR6+, CD45RA+, CCR7- |
| CD4 Th1_17 EMRA | CD3+, CD4+, CXCR3+, CCR6+, CD45RA+, CCR7- |
| CD8_Naive | CD3+, CD8+, CD45RA+, CCR7+ |
| CD8_CM | CD3+, CD8+, CD45RA-, CCR7+ |
| CD8_EM | CD3+, CD8+, CD45RA-, CCR7- |
| CD8_EMRA | CD3+, CD8+, CD45RA+, CCR7- |

Supplementary figure 2: ROC curves for diagnostic classification contrasting principal component scores to plasma neurodegenerative biomarkers

AD:

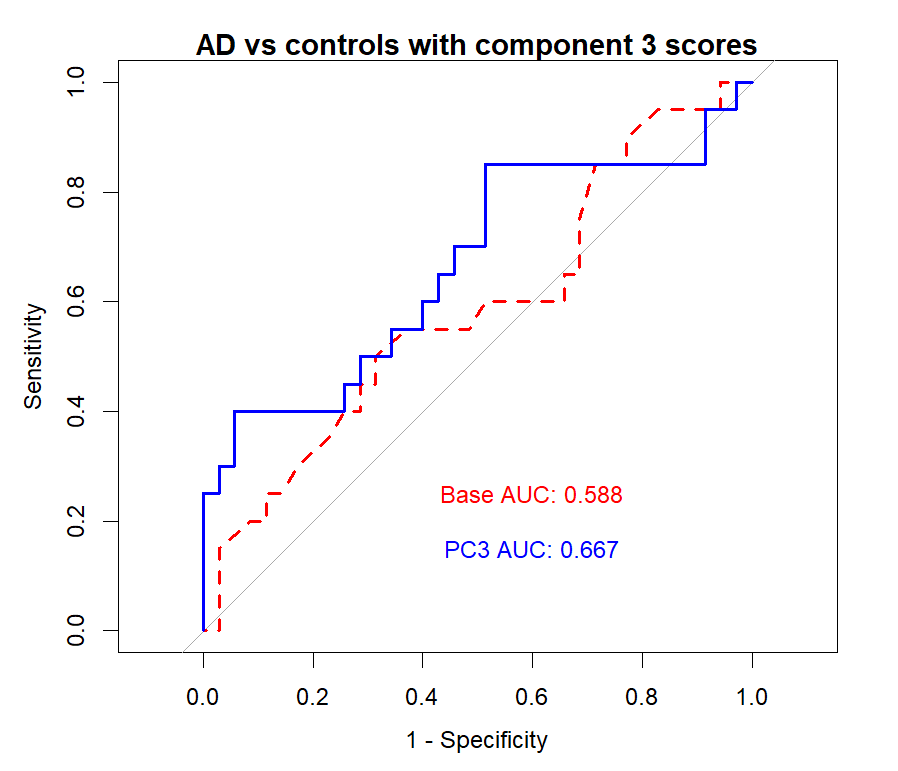

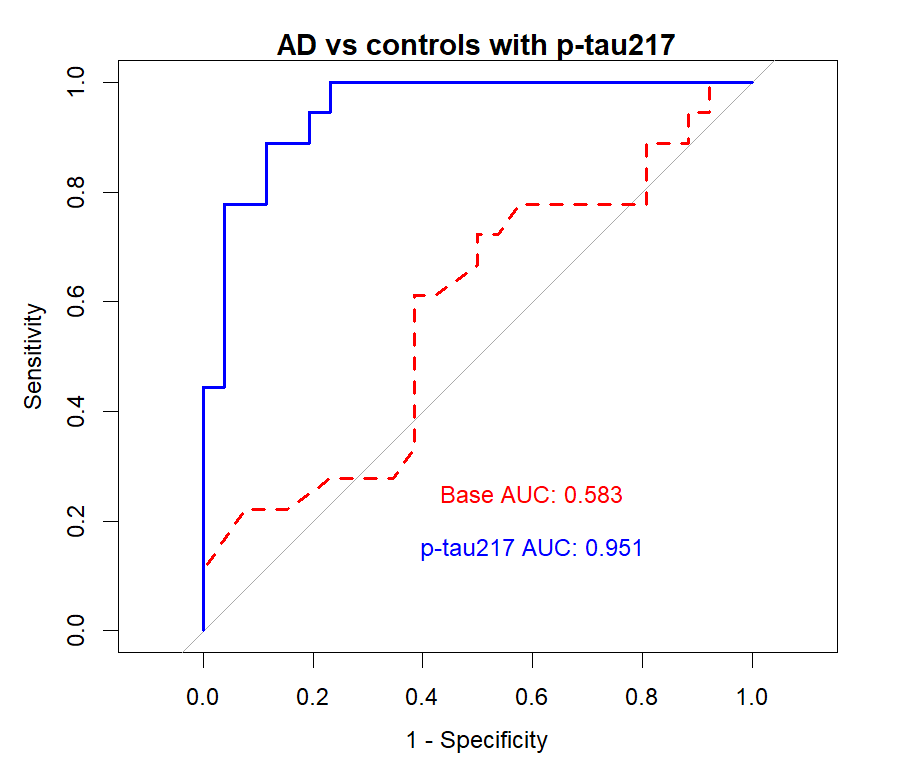

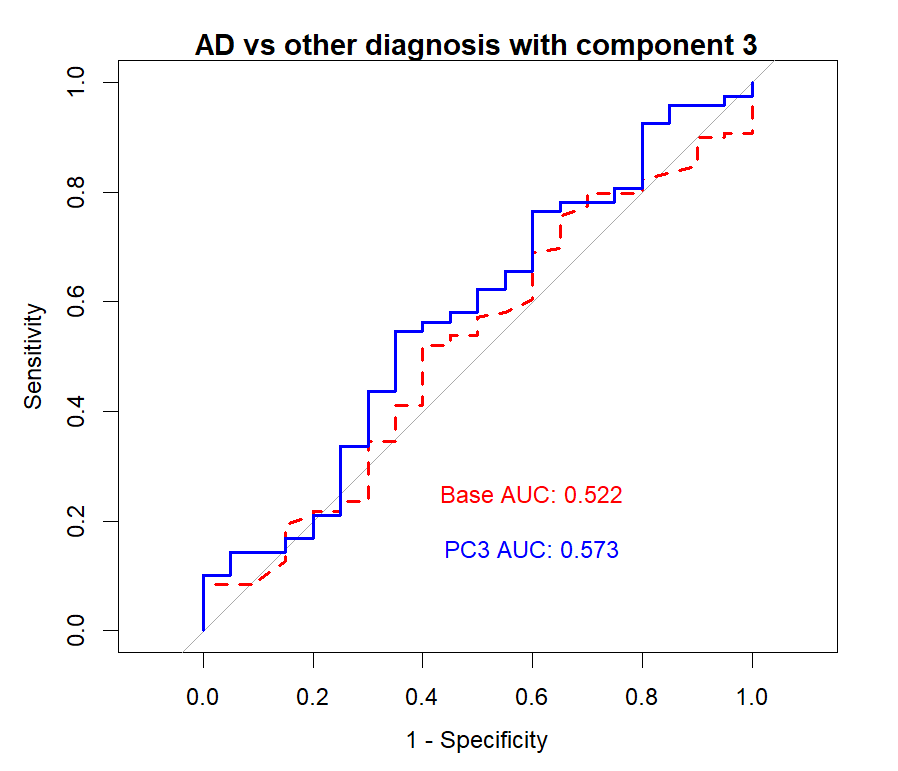

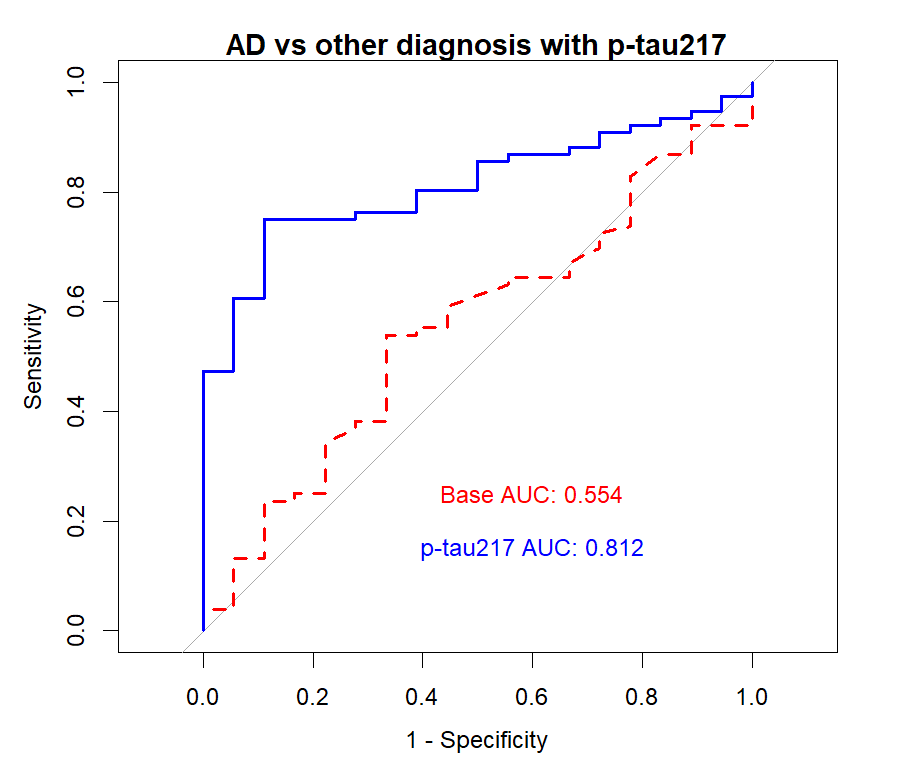

FTD:

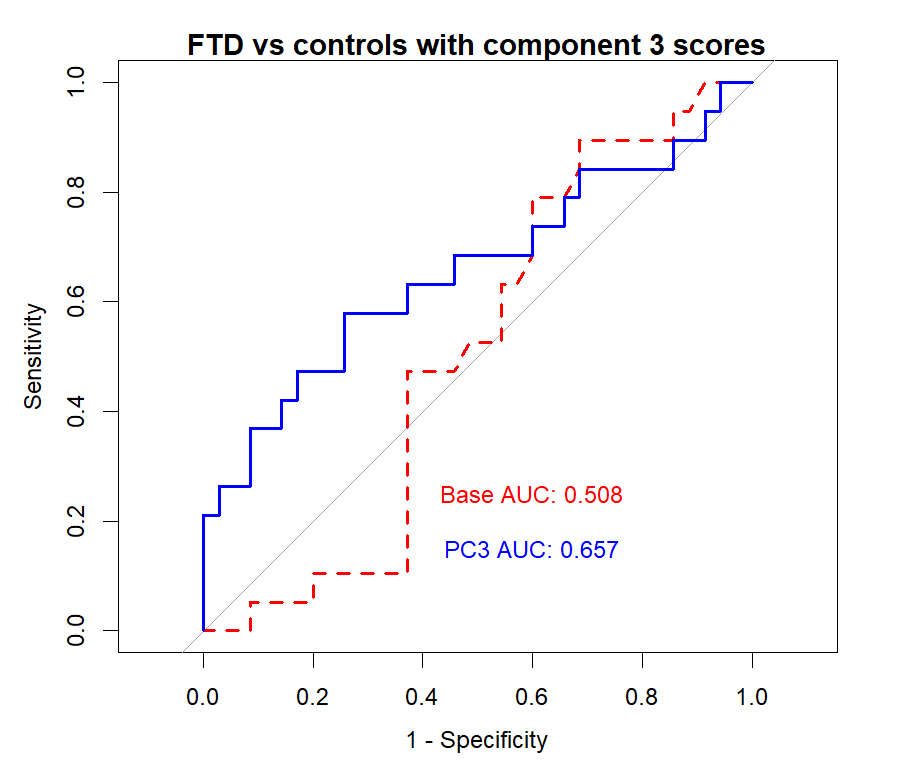

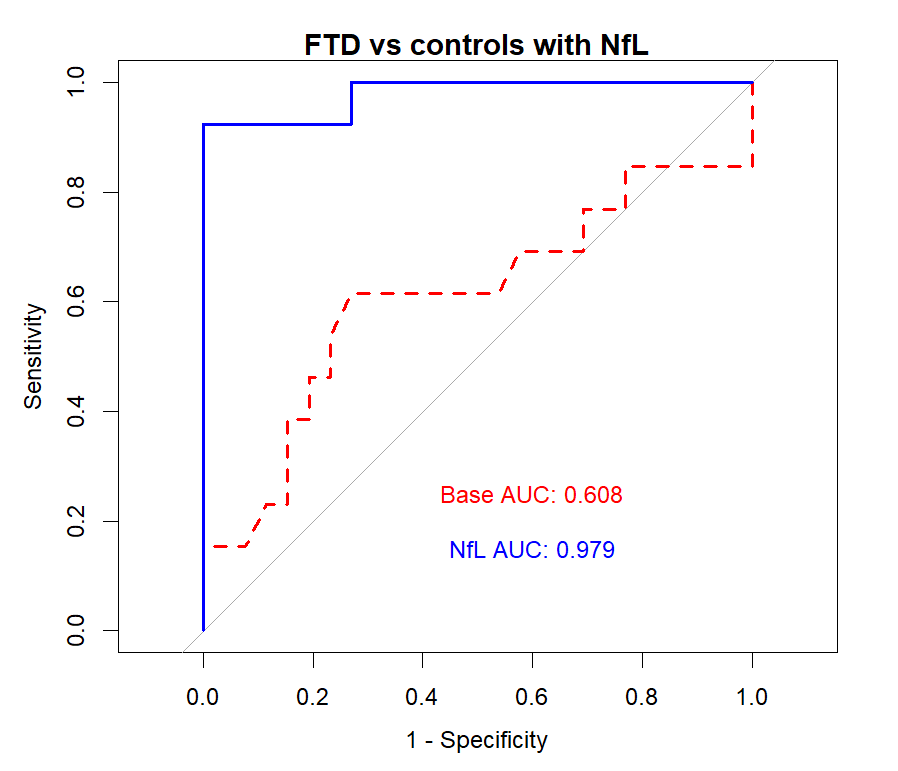

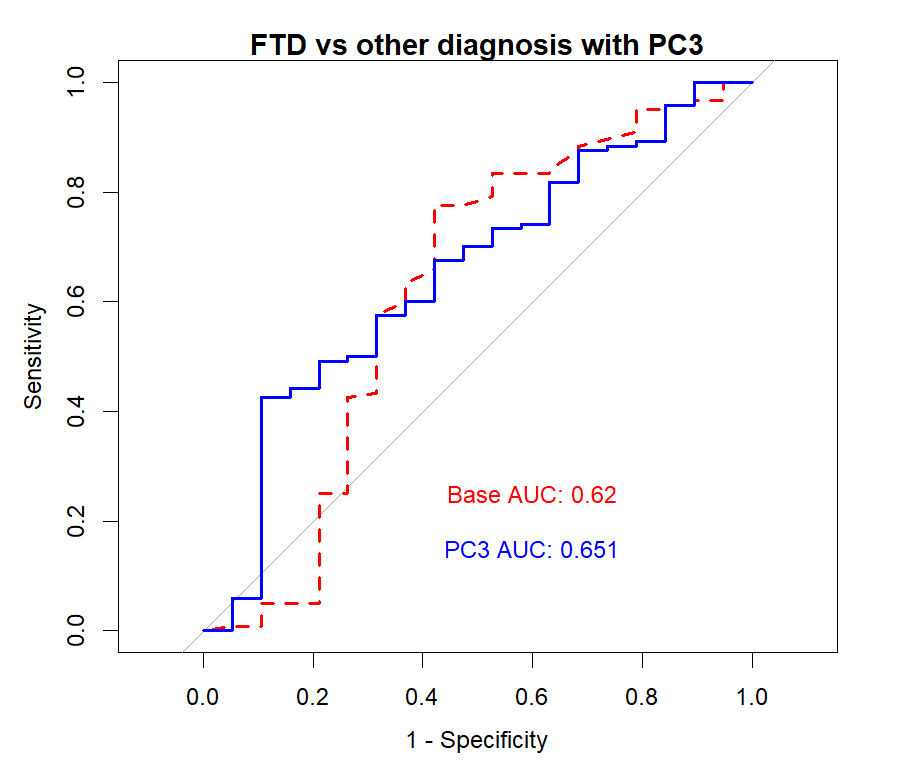

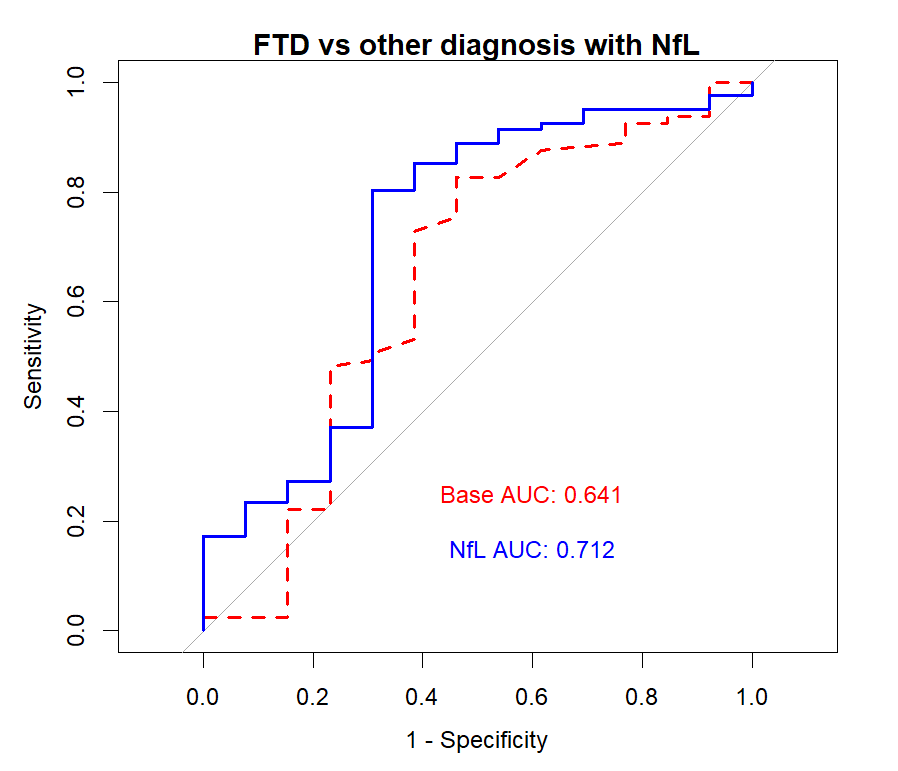

Supplementary figure 2: Top panel: Using component 3 scores, there was a significant improvement in the classification of AD compared to controls from the base model including covariates (AUC=0.59 to AUC=0.66, p=0.036), but again this was limited in comparison to the inclusion of p-tau217 in the model which improved the classification from AUC= 0.58 to AUC=0.95 (p<0.001). There was no significant improvement in classification of AD compared to other diagnostic group (AUC=0.52 to AUC=0.57, p=0.25); in contrast, addition of p-tau217 improved classification from AUC=0.55 to AUC=0.81 (p=0.001).

Bottom panel: Including component 3 scores improved the classification of FTD vs controls (AUC=0.51 to AUC=0.66, p=0.022), that although statistically significant provided very little improvement compared to a model with NfL (AUC=0.61 to AUC=0.98, p<0.001). There was no significant improvement in classification between FTD and other diagnostic groups (AUC=0.62 to AUC 0.65, p=0.15), whilst there was a significant improvement in classification with NfL (AUC=0.64 to AUC=0.71, p=0.046).
